# Validation of a glycomics-based test associated with risk of HCC development in cirrhosis

**DOI:** 10.1101/2024.02.27.24303387

**Authors:** Xavier Verhelst, Leander Meuris, Roos Colman, Anja Geerts, Annelies Van Hecke, Hans Van Vlierberghe, Nico Callewaert

**Affiliations:** Ghent Liver Research Center, Ghent University, Ghent, Belgium; Department of Hepatology and Gastroenterology, Ghent University Hospital, Ghent, Belgium; VIB-Ugent Center for Medical Biotechnology, Technologiepark-Zwijnaarde 75, B-9052 Ghent, Belgium; Department of Biochemistry and Microbiology, Ghent University, Ledeganckstraat 35, B-9000 Ghent, Belgium; Biostatistics Unit, Department of Public Health and Primary Care, Ghent University, Ghent, Belgium

**Author notes:** Corresponding author **Correspondence should be addressed to** Xavier Verhelst, Ghent University Hospital, Corneel Heymanslaan 10, Route 1241, Ghent B-9000, Belgium. Authors contributed equally (Xavier Verhelst, Leander Meuris). Authors contributed equally (Hans Van Vlierberghe, Nico Callewaert). **Email addresses**, Leander Meuris, Roos Colman, Anja Geerts, Annelies Van Hecke, Hans Van Vlierberghe, Nico Callewaert.

**Keywords:** Glycomics, glycocirrhotest, hepatocellular carcinoma (HCC), cirrhosis, prognostic biomarker, risk stratification

## Abstract

**Background:** Cirrhosis is the main risk factor for the development of Hepatocellular carcinoma (HCC). Six-monthly screening with ultrasound is advocated for the surveillance of cirrhotic patients. We recently showed that a glycomics-based test (GlycoCirrhoTest [GCT]) can provide additional information regarding the risk of HCC development in cirrhotic patients.

**Aims:** Independent clinical validation of the GCT for the assessment of the risk of HCC development in cirrhosis and exploration of additional clinical parameters to assess HCC risk.

**Methods:** Validation study on serum samples of patients with established compensated cirrhosis (CHILD Pugh A & B) in a tertiary liver centre. Serum N-glycan profiling was performed and GCT was calculated at baseline. During the follow up period, patients were screened for the presence of HCC every 6 months with ultrasound.

**Results:** A total of 198 cirrhotic patients were followed in clinical routine for the development of HCC. 29 patients developed HCC and one died during follow up. At baseline, the mean GCT value was significantly higher in patients who developed HCC within 3 years compared to patients who did not develop HCC (Welch’s t-test, p-value 3 years: 0.034). A high GCT at baseline was associated with increased HCC incidence with a HR of 5.8 (95% CI: 0.7 – 48), 4.8 (95% CI: 1.4 – 16) and 3.6 (95% CI: 1.2 – 11) at 3, 5 and 7 years post sampling respectively. Results from this study are in agreement with previous results^1^, as shown in a meta-analysis. Moreover, we also identified albumin as an independent predictor for developing HCC in a multivariate analysis revealing that low albumin blood levels (< 4g/dL) are also associated with increased HCC incidence with a HR at 7 years of 2.3 (95% CI: 1.1 - 4.9). For subjects with both high GCT and low albumin we found a HR of 9.8 (95% CI: 3.5 to 27) at 7 years.

**Conclusions:** GCT is a glycomics-based test that provides additional information for risk assessment of HCC development in cirrhosis. This information could be used to develop personalised HCC screening programs in cirrhotic patients according to the value of GCT. Serum albumin levels could provide additional and GCT-independent information which may add to the utility of the test.

## Introduction

Hepatocellular carcinoma (HCC) is the most common type of primary liver cancer and represents a major health burden^2^. Globally, the presence of cirrhosis is the most important risk factor for the development of HCC. Consequently, HCC should be a preventable disease, since regular screening for HCC in patients with cirrhosis should lead to an early stage diagnosis, at a time when the patient is still amenable to curative treatment options. For this reason, for decades the practice of screening for HCC in patients with cirrhosis has been advocated by authoritative professional associations such as EASL and AASLD. Unfortunately, clinical reality is more complicated and adherence to screening regimens is disappointingly low. In a recent study including more than 82000 patients with cirrhosis in the USA, only 8.78% received HCC surveillance^3^, a finding in line with older reports^4^. These low rates are explained mainly by patient- and physician-related factors and geographic variation. Recently, the evidence for HCC surveillance in cirrhosis was questioned due to the lack of randomised clinical trials showing its benefits^5^.

Today, HCC screening is a ‘one-size-fits-all’ approach. However, cirrhosis comes in different stages and patients develop cirrhosis due to different underlying liver diseases. HCC incidence rates can vary with the primary aetiology of liver disease^6^ and with disease stage (e.g. HBsAg clearance in Hepatitis B virus infection)^7^. Other (independent) risk factors such as sex, ethnicity, age, tobacco use and many more have also been associated with the development of HCC in various disease settings. In an ideal world, a successful screening strategy would take into account such independent risk factors, as well as the primary disease aetiology to determine whether a patient is at high or low risk of developing HCC. To this end, EASL has recently published a policy statement^8^, advocating the stratification of cirrhosis patients into low, medium and high risk groups. Low risk patients would not receive any screening, medium risk patients would receive standard ultrasound screening and high risk patients would receive MRI-based screening, which has much higher specificity and also a higher sensitivity when compared to standard ultrasound. However, there is currently no consensus on how to stratify patients. Moreover, necessary information can be incomplete or lacking, which further complicates the matter (e.g. information pertaining to socially sensitive issues such as drinking behaviour).

Molecular prognostic markers that provide information about an individual’s risk of developing HCC and which can be assessed non-invasively, could help address the issue of patient stratification in a more direct way. Fujiwara *et al*. and Marasco *et al*. extensively reviewed clinical risk scores and biomarkers allowing for a more granular assessment of the risk of HCC development in cirrhosis patients^9,10^. Most clinically accepted risk markers are based on routinely measurable parameters, such as bilirubin, platelets, albumin, AST, … possibly combined with sex, age, BMI, aetiology or other patient-related data. Examples of this are the aMAP risk score^11^, the aspartate aminotransferase to platelet ratio index (APRI) and the Fibrosis-4 index (Fib-4). Another clinically accepted approach is the use of liver stiffness measurements (LSM), for which a number of large studies have shown utility in HCC risk assessment, mostly in viral hepatitis cohorts^12–14^. Other, less established but potentially interesting HCC risk markers include serum proteins^15,16^ but also genomic^17^, transcriptomic^18^ and glycomic^1^ signatures that shed light on the underlying pathophysiology of carcinogenesis and thus the related risk of HCC development. However, none of these have found widespread use in the clinic until today.

Over the last decade or two, the value of serum glycomics as a source of biomarker development in liver disease has become clear^1,19–23^. Previously, our teams showed that a ‘glycomics’ biomarker based on the total serum N-glycan profile, could distinguish chronic liver disease patients with established but compensated cirrhosis from those with earlier stages of fibrosis^23^. This biomarker, also called GlycoCirrhoTest (GCT), is not only diagnostic for the presence of cirrhosis, but is associated with the risk of development of HCC in cirrhotic patients. We observed significantly increased baseline values of GCT in patients with cirrhosis who developed HCC after a median follow-up of 6.4 years as compared with patients who did not^1^. Using an optimised cut-off of 0.2, the hazard ratio (HR) for HCC development over the entire study was 5.1 [95% confidence interval (CI), 2.2–11.7; p < 0.001], and the HR for HCC development within 7 years was 12.1 (95% CI, 2.8–51.6; p < 0.01).

GCT is characterized by a relative increase in bisecting N-acetylglucosamine (GlcNAc)–containing N-glycans and a relative decrease in triantennary N-glycans on glycoproteins in serum. The enzyme responsible for the biosynthesis of bisecting GlcNAc residues on N-glycans is N-acetylglucosaminyltransferase III (GnT-III)^24^. In rat models, bisecting GlcNAc residues or GnT-III enzyme activity are not detectable in healthy liver tissue^24,25^. However, in rats exposed to 1,2-dimethylhydrazine or diethylnitrosamine, GnT-III is expressed at significant levels in cirrhotic nodules. Significantly increased GnT-III activity has been observed in serum and liver tissue of patients with nodular cirrhosis and HCC but not in patients with chronic hepatitis without cirrhosis^26,27^. This evidence points towards a role for this glycomics biomarker as a surrogate marker of increased nodular regenerative activity in cirrhosis, associated with an observed increased risk of HCC formation.

The goal of this study is to provide an independent external validation of this glycomics biomarker as a risk stratification tool for HCC risk in cirrhosis.

## Patients and methods

### Study cohort and study design

We used serum samples of 341 patients with cirrhosis from the biobank of the department of Gastroenterology and Hepatology at Ghent University Hospital (Belgium). After exclusion of duplicate database entries, subjects with current or previous HCC, and subjects with Child-Pugh class C, 198 subjects were selected and monitored for the development of HCC during routine follow-up (Figure 1). All patients received screening for HCC using liver ultrasound and AFP measurements every 6 months. Upon suspicion of HCC on ultrasound, a confirmation with MRI was performed. Follow-up was upon HCC diagnosis, liver transplantation or patient death. Demographic and clinical data are summarised in table 1.

**Table 1.** Baseline characteristics.

|  | All patients | No HCC | HCC | P (Fisher exact test) |
| --- | --- | --- | --- | --- |
| Sex (M/F) | 138/60 (69.7%/30.3%) | 115/54 (68.0%/32.0%) | 23/6 (79.3%/20.7%) | 0.278 |
| Etiology |  |  |  |  |
| Alcohol | 92 | 78 (46.2%) | 14 (48.3%) | 0.645 |
| HCV | 31 | 24 (14.2%) | 7 (24.1%) |  |
| HBV | 23 | 21 (12.4%) | 2 (6.9%) |  |
| Mixed | 16 | 15 (8.9%) | 1 (3.4%) |  |
| AIH | 14 | 13 (7.7%) | 1 (3.4%) |  |
| Other | 22 | 18 (10.7%) | 4 (13.8%) |  |
|  | Mean (±SD) | Mean (±SD) | Mean (±SD) | P (Mann-Whitney test) |
| Age | 55.8 (11.6) | 55.4 (11.8) | 57.9 (10.6) | 0.220 |
| Time at risk (years) | 7 (4.1) | 7.3 (4.1) | 5.6 (3.7) | **0.046** |
| Total bilirubin (mg/dL) | 1.4 (1.5) | 1.4 (1.6) | 1.1 (0.7) | 0.836 |
| Albumin (g/L) | 41 (6) | 41 (6) | 40 (7) | 0.208 |
| Creatinine (mg/dL) | 1.0 (0.5) | 1.0 (0.6) | 0.9 (0.2) | 0.178 |
| AST (U/L) | 48.6 (32.9) | 47.5 (33.1) | 54.4 (31.7) | 0.163 |
| ALT (U/L) | 43.6 (42.9) | 42.7 (44.2) | 48.0 (36.2) | 0.082 |
| Platelets (10<sup>9</sup>/L) | 148.3 (80.7) | 151.1 (80.5) | 133.5 (81.7) | 0.207 |
| INR | 1.2 (0.3) | 1.2 (0.3) | 1.2 (0.1) | 0.791 |
| Child-Pugh | 5.7 (1.1) | 5.7 (1.1) | 5.4 (0.9) | 0.133 |

**Figure 1.**
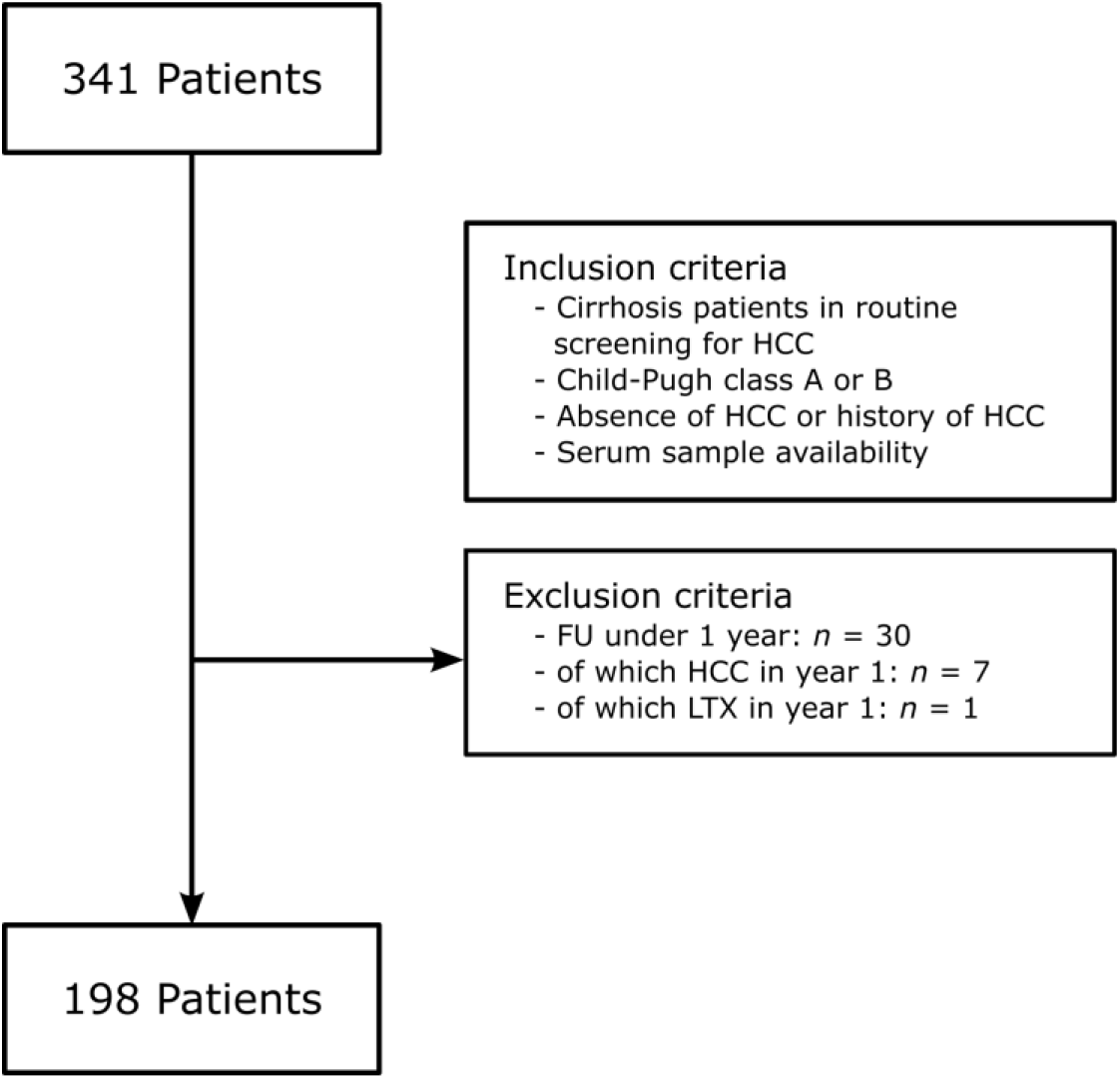
Inclusion and exclusion of patients.

Serum samples were collected at the start of clinical follow up through classic venepuncture directly in serum preparation tubes (BD vacutainer SST tubes with silica clot activator and polymer gel). Serum was prepared according to the manufacturer’s instructions. A part was subjected to standard clinical biology procedures and another part aliquoted into standard Eppendorf tubes and immediately stored at -20°C or -80°C until the time of glycomics analysis. For the meta-analysis, we cleaned and the reused the data from a cohort that we previously published^1^.

### Glycomics analysis

The serum N-glycan profiles were determined as previously described^28^. In summary, N-glycans were prepared using the ‘on-membrane’ protocol. Three μl of serum was denatured and the proteins bound to a polyvinylidene fluoride (PVDF) membrane. The denatured proteins were reduced and carboxymethylated and N-glycans were released overnight at 37°C with 5 mU (IUBMB) of PNGase F. The following day, the N-glycan samples were collected in a new vial and dried in a SpeedVac at 37°C, followed by overnight labelling at 37°C with 1 μl of labelling solution (a 1:1 [vol/vol] mixture of 20 mM APTS in 1.2 M citric acid and 750 mM picoline borane in dimethyl sulfoxide [DMSO]). The following day, the glycans were cleaned up over a Sephadex G10 resin and, after elution, subjected to exoglycosidase treatment with *A. ureafaciens* sialidase (in house production). The desialylated glycans were then analysed by capillary gel electrophoresis on an ABI 3500 genetic analyser equipped with a 50 cm 8-capillary array. We used POP7 polymer to fill the capillaries and 100 mM TAPS buffer pH 8.0 containing 1 mM EDTA as the cathode and anode buffer. Electrokinetic injection of the glycan samples was performed for either 5 or 10 seconds at 15 kV, depending on the signal strength for each sample.

### Statistical analysis

Statistical analysis was performed using R 4.2^29^. Summary statistics were calculated using the summarytools package^30^. Time at risk was defined as the follow-up time until development of HCC, death or end of follow-up without HCC development. For the Mann-Whitney U tests we used the function from the coin package^31^ with asymptotic approximation of the null.

For survival analysis, an event was defined as a new diagnosis of HCC in patients not previously diagnosed with HCC. After applying the exclusion criteria, the dataset (*n* = 198) contained one patient who died during their FU, and we considered this to be an event as well. Survival analysis was performed with the survival^32^ and rms^33^ packages and visualisations with the survminer^34^ package. Log-rank tests and hazard ratios at 3, 5 and 7 years and their 95% confidence intervals were calculated based on Cox proportional hazards regression. We assessed the proportional hazards (PH) assumption in the Cox models both graphically (complementary log-log survival versus log time) and via the score test that is available in the survival package. When the PH assumption was not fulfilled or the results were doubtful we used a stratified Cox model for those covariates. Model building followed a step up strategy and decisions were based on likelihood ratio testing.

To generate the leaf plots, we first calculated combinations of sensitivity and specificity for different test cut-offs with the ROCit package^35^. Since the number of events was not very high, especially at the 3 year time point, we used the nonparametric ROC curve estimation method, which applies a kernel-based smoothing of the empirical ROC curve. Such an approach results in conservative AUC values, but avoids large jumps in sensitivity and specificity estimates at different test cut-off values in the case of a (very) discrete empirical ROC curve. We then retrieved the specificity and sensitivity values associated with test cut-offs of 0.15, 0.20 and 0.25 at 3, 5 and 7 years and used those as input for the leaf plots. The script for producing the leaf plots was adapted from a script by Zampieri and Einav^36^.

### Ethics

The study was approved by the ethical review board of Ghent University Hospital. Informed consent was obtained from all patients. The study complies with the requirements of the Declaration of Helsinki.

## Results

### Baseline characteristics

Serum N-glycomics profiles were obtained in 228 patients with cirrhosis and the value of GCT was calculated. Child-Pugh C patients and patients who developed HCC during the first year of follow up were excluded, similar to the design of the pilot study^1^. We refer to figure 1 for an overview of the inclusion and exclusion criteria. 198 patients were included in the final analysis (Child-Pugh A; n=162 (81.8%); Child-Pugh B; n=36 (18.2%)). After a mean time at risk of 7 years, 29 patients developed HCC and one died. In this cohort, no patients received liver transplantation. Baseline characteristics (table 1) were comparable between patients who developed HCC and those who did not, with the exception of the mean time at risk, which was longer in the no HCC group (Mann-Whitney U test p-value: 0.046).

### GCT is a dynamic biomarker associated with the development of HCC

Applying the same cut-off for GCT as in our proof-of-concept study^1^ (cut-off = 0.2), 111 patients (56.1%) showed a GCT value above this cut-off and 87 patients (43.9%) below this value. From figure 2, it becomes clear that the mean GCT value, taken at baseline, is significantly higher in patients who developed HCC within 1 to 3 years after taking the measurement compared to patients who did not develop HCC within that timeframe (Welch’s t-test, p-value: 0.030). The difference is smaller and not significant anymore (Welch’s t-test, p-value: 0.258) for patients who only developed HCC later, within 3 to 7 years after the baseline GCT measurement. Figure 2 shows the distribution of GCT values in both cases.

**Figure 2.**
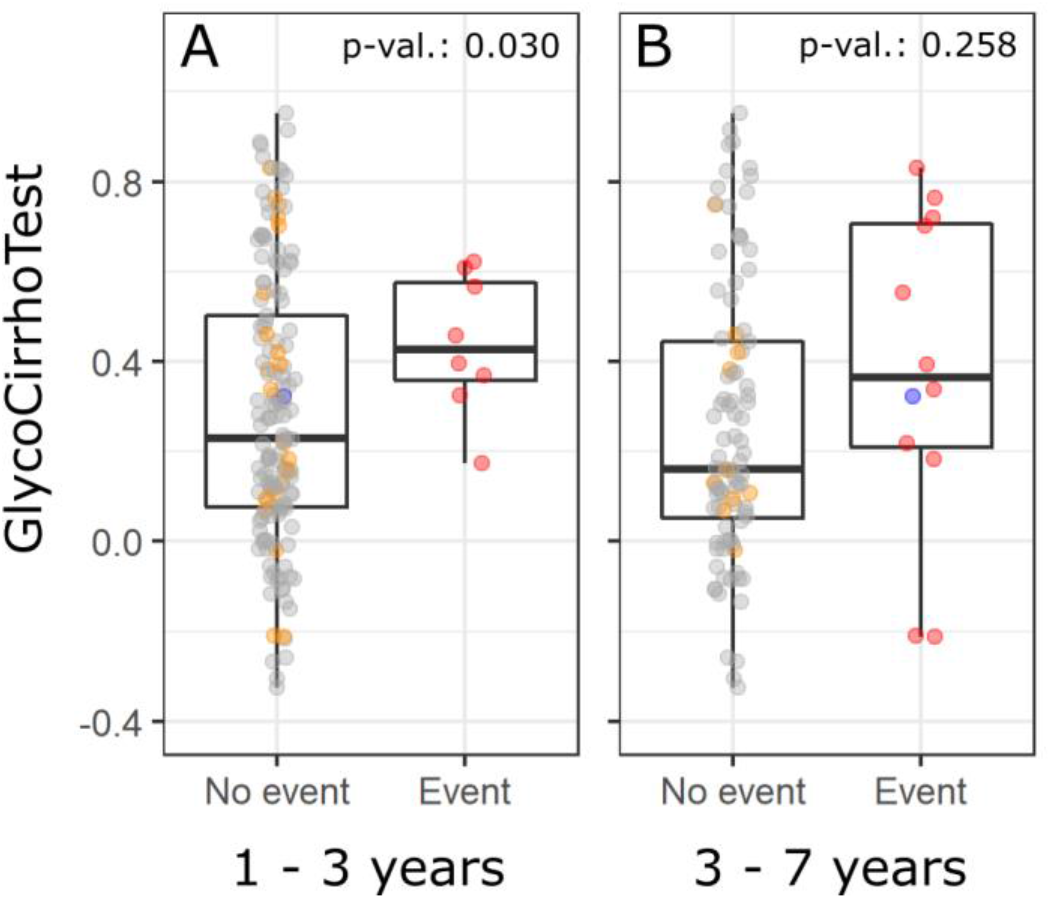
Distribution of the GCT value in groups at different times after sampling. The ‘No event’ group consists of subjects who had a follow up time of at least 3 years (panel A) or at least 7 years (panel B) without an event. Within this group, there are subjects who never got an event during their follow up (grey dots), did develop HCC at some point during their follow up after more than 3 resp. 7 years (orange dots) or who died later in the study (blue dot). The ‘Event’ group consists of subjects who developed HCC (red dots) or died (blue dot) within the indicated time frame below the plot. The mean GlycoCirrhoTest value was significantly different between no event and event groups for the 0 - 3 years time frame but not for the 3 - 7 years time frame (Welch’s t-test, p-value 0 - 3 years: 0.030, 3 - 7 years: 0.258).

Interestingly, patients that developed HCC earlier thus seem to have a generally higher baseline GCT value but we also see that after a longer period, the test seems to be less informative. Several patients developed HCC after longer than 7 years of follow-up. GCT values are also not informative after such a long follow-up period (data not shown).

### Value of GCT as a prognostic marker of HCC

Cumulative incidence curves (figure 3) clearly illustrate the discriminative ability of GCT for risk assessment of HCC development. The hazard ratio with a 95% confidence interval is shown at 3, 5 and 7 years, along with log-rank test p-values. The best predictive power seems to be at 4-5 years. The HR at 5 years is 2.9 (95% CI: 1.2 to 7.0). From figure 3, it is also clear that in the group of patients with a GCT value below 0.2, there is only one case of HCC within the first 3 years after sampling, while in the group with high GCT values, HCC cases start to arise soon after the start of sampling. In this cohort of mainly alcoholic cirrhosis, HCC incidence is lower than in the discovery cohort with almost exclusively viral hepatitis^1^, which is in line with earlier findings.

**Figure 3.**
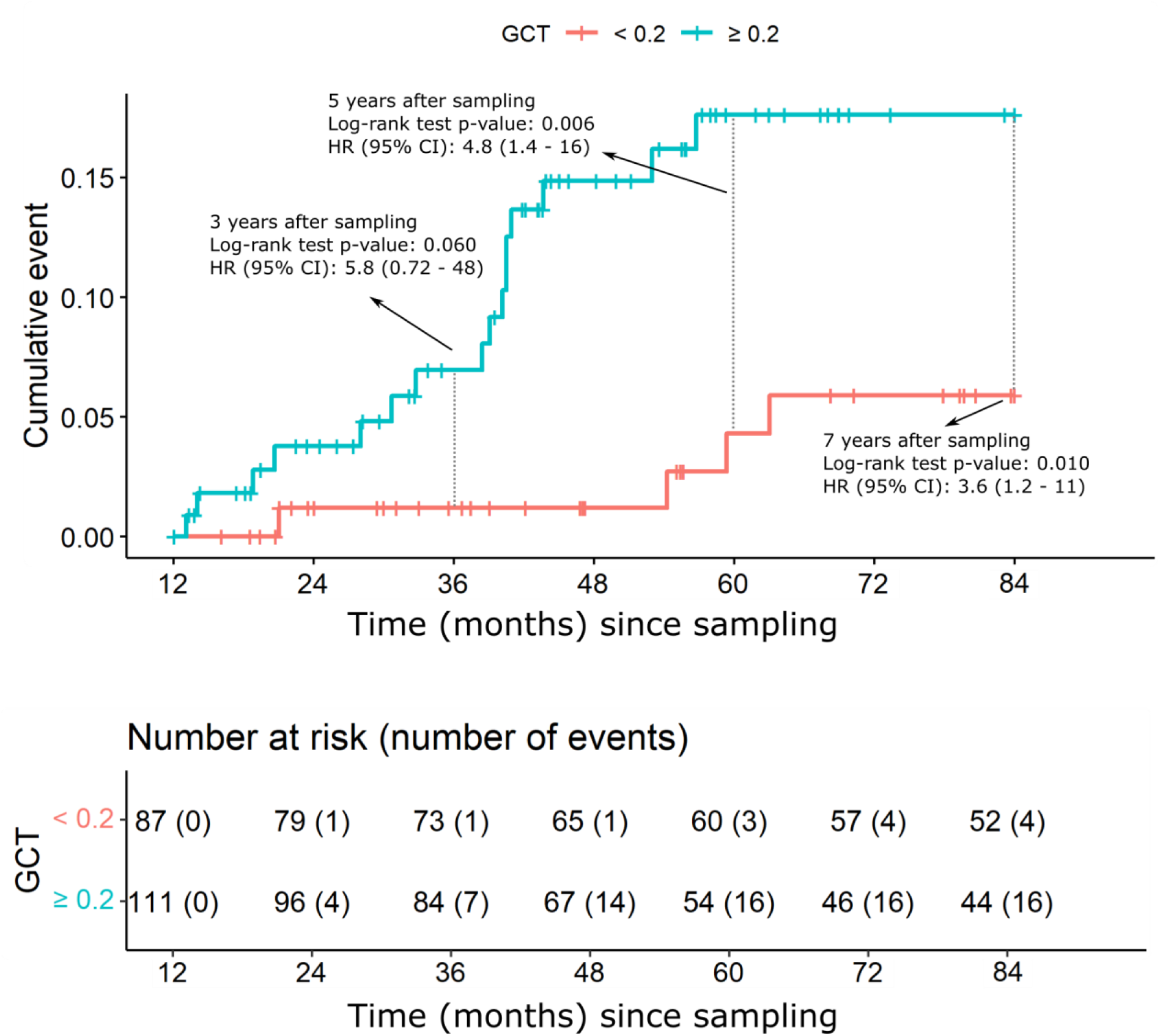
Estimated cumulative incidences for HCC according to the value of the GlycoCirrhoTest. Red line GCT < 0.2 at time = 0; blue line GCT ≥ 0.2 at time = 0. The log-rank test p-values and hazard ratios (HR) along with a 95% confidence interval for the HR are also indicated for the 3, 5 and 7 years time points. Below, the number of events and subjects at risk in each GCT group at the indicated times are given.

In order to fully appreciate the clinical utility of this prognostic test, leaf plots^37^ were designed for the assessment of risk of HCC development at 3, 5 and 7 years, using different test cut-off levels (figure 4). These confirm the high negative predictive value of this biomarker. For example, when we put forward a naive estimate of 5 % for the pre-test probability of developing HCC within the first 3 years and using a test cut-off of 0.2, then the post-test probability of developing HCC is 1.1 % given a negative test. This corresponds to a negative predictive value (NPV) of 98.9%. Table 2 lists the negative test likelihood ratios for different test cut-off values and follow up periods and the associated NPV values given a specified pre-test probability of developing HCC within the given time period. It is clear from both figure 4 and table 2 that the test has most clinical utility when a negative test is used to rule out future HCC within 3 years after the test, as represented by a relatively larger green area in figure 4 and higher NPV in table 2. A lower cut off (more ‘stringent’ from the point of view of ruling out high risk) results in a better stratification. Of course this is a trade-off since a more stringent test restricts making decisions to fewer subjects. With a test cut-off of 0.2 (figure 4, middle panels), which is just below the median GCT value for the ‘no HCC’ group, we can confidently stratify about 45% of the subjects under surveillance as low HCC risk subjects. Patients who receive a negative test at baseline have a 5 times lower probability of developing HCC within 3 years after sampling.

**Table 2.** Illustration of clinical utility of the GlycoCirrhoTest to rule out future HCC.

| Cut-off | Years post sampling | Negative test LR | Pre-test prob. of HCC | Post-test prob. of HCC (neg. test) | NPV |
| --- | --- | --- | --- | --- | --- |
| 0.15 | 3 | 0.146 | 5.0 % | 0.8 % | 99.2 % |
| 0.20 | 3 | 0.208 | 5.0 % | 1.1 % | 98.9 % |
| 0.25 | 3 | 0.289 | 5.0 % | 1.5 % | 98.5 % |
| 0.15 | 5 | 0.324 | 10.0 % | 3.5 % | 96.5 % |
| 0.20 | 5 | 0.373 | 10.0 % | 4.0 % | 96.0 % |
| 0.25 | 5 | 0.438 | 10.0 % | 4.6 % | 95.4 % |
| 0.15 | 7 | 0.459 | 15.0 % | 7.5 % | 92.5 % |
| 0.20 | 7 | 0.483 | 15.0 % | 7.9 % | 92.1 % |
| 0.25 | 7 | 0.526 | 15.0 % | 8.5 % | 91.5 % |

**Figure 4.**
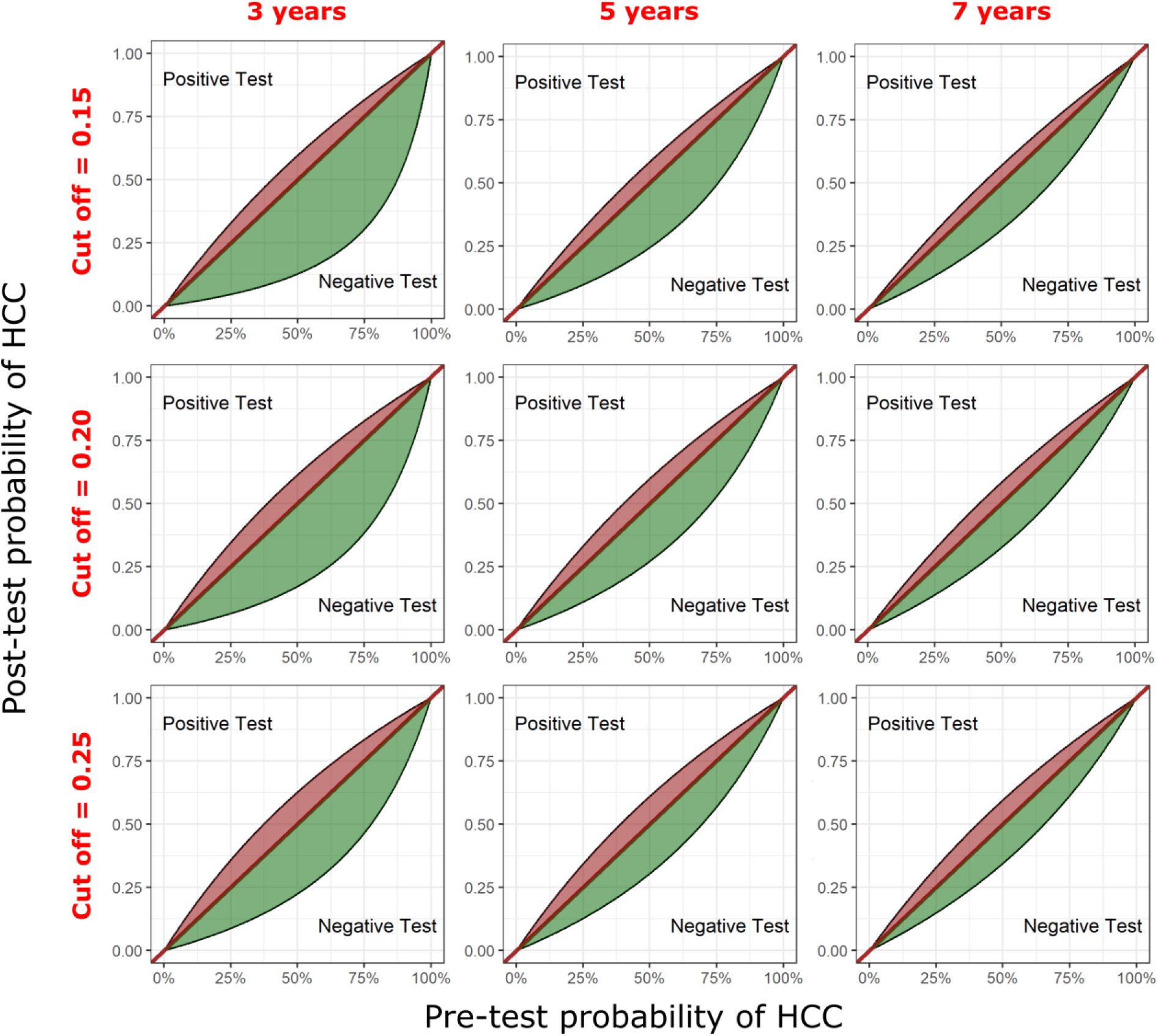
Leaf plots for different GCT test cut-off values and for 3, 5 and 7 years after sampling. The plots show the post-test probability of developing HCC when the test is negative or positive (curved black lines) depending on the pre-test probability of developing HCC. A larger green-shaded area indicates that the test is most informative when it turns out negative and used to rule out high risk of developing HCC. Small red shaded areas show that the test is less informative when it turns out positive and should thus not be used to rule in subjects at high risk of developing HCC.

### Meta-analysis and exploration of covariate effects

To assess the agreement between this study and the proof-of-concept study^1^, we performed a meta-analysis. The results are shown in figure 5. In both studies, we found that high GCT test values (cut off 0.2) were associated with similarly increased HCC hazard rates. We found comparable hazard ratios at 3, 5 and 7 years post sampling (Figure 5). Individual and overall hazard ratio’s for developing HCC in the high GCT versus low GCT group are significantly increased at 5 and 7 years post sampling but not yet at 3 years. Both studies are in agreement with respect to this finding. Finally, we found a pooled hazard ratio of 3.82 (95% CI: 1.67 to 8.75) and 4.86 (95% CI: 2.27 to 10.45) at 5 and 7 years respectively (Figure 5).

**Figure 5.**
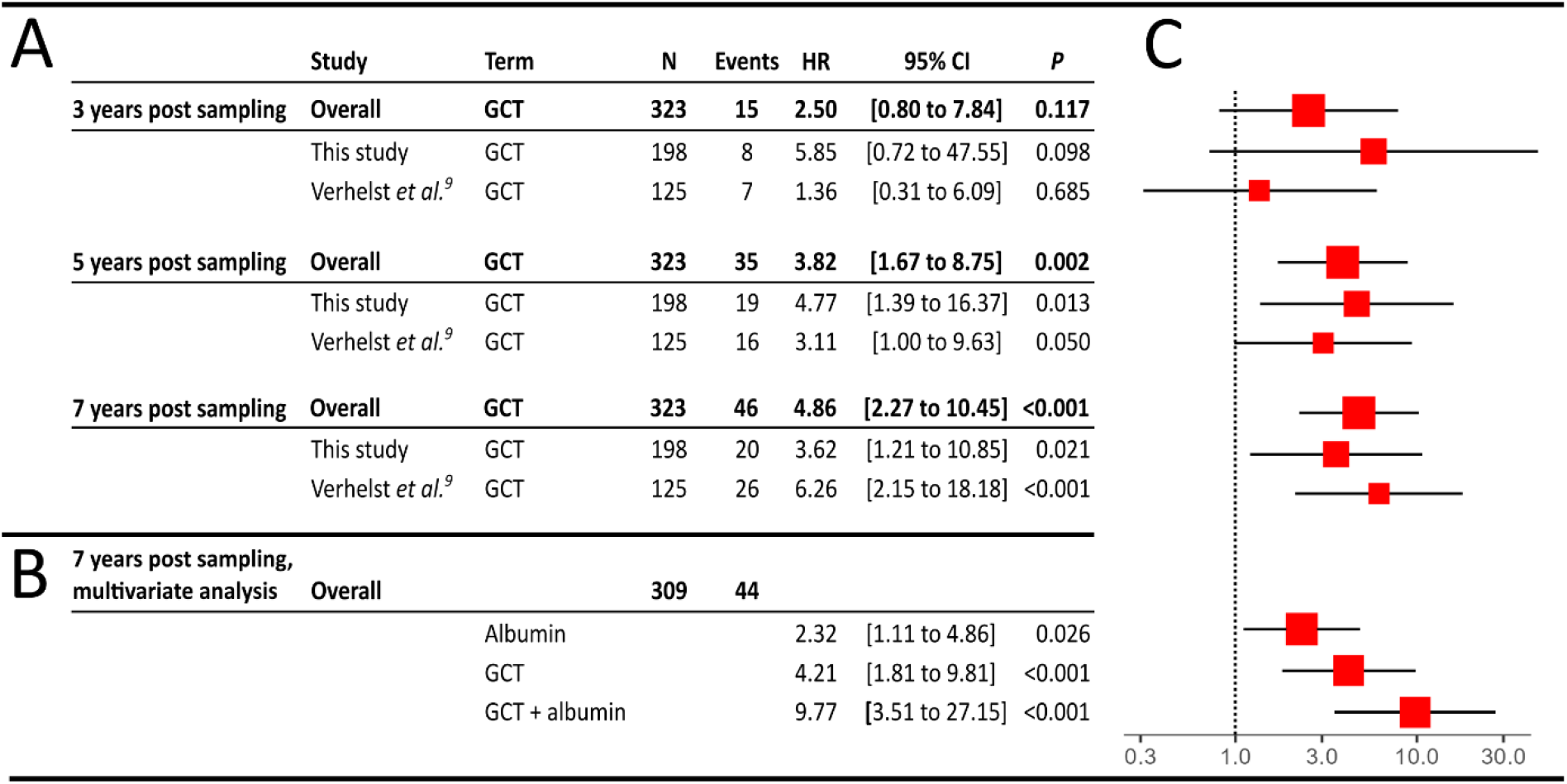
Meta-analysis of this study and Verhelst *et al*. (2017)^1^ and covariate effect exploration. The hazard ratio for GCT (cut off 0.2) in each study and at 3, 5 or 7 years after sampling is reported (panel A). The results of the multivariate analysis at 7 years are also shown (panel B). For each scenario, the corresponding 95% confidence intervals are shown with the p-values indicating whether the HR is significantly different from 1. The same information is also provided graphically (panel C), HR estimates are given as red squares, the size being proportional to the cohort size. 95% confidence intervals are given as black lines.

Combining the data from both studies resulted in a dataset with a sufficient number of events at 7 years to assess whether other clinical variables may modulate the GCT signal or independently predict HCC risk. We built a Cox proportional hazards model adding primary etiology (alcohol, HBV, HCV or other, the latter covering all other etiologies of chronic liver disease), cohort (this study versus Verhelst *et al*.^1^) and albumin to the model. Albumin was dichotomized with lower than or equal to 4 g/dL serum albumin being the cut off. This cut off is based on our own observations but also corresponds to a previously published report stating that serum albumin concentrations below 4 g/dL are predictive of a high HCC risk in HCV patients^38^. During model building, we found that the PH assumption was not valid for cohort and etiology. Consequently, we included these as strata in a stratified Cox model, which allows to adjust for their effects. We found no evidence for interaction effects between any of the four covariates. In our final model, we thus found that high GCT or low albumin were independently predictive of a higher HCC risk at 7 years after sampling, with a HR of 4.2 (95% CI: 1.8 to 9.8) for GCT and a HR of 2.3 (95% CI: 1.1 to 4.9) for albumin. When a patient has both a high GCT and low albumin, the estimated HR is 9.8 (95% CI: 3.5 tot 27.1) compared to patients that have a low GCT and normal albumin. Since we found no evidence for interaction effects with cohort or etiology, these HRs are assumed to be the same regardless of the cohort or etiology.

## Discussion

Early diagnosis of HCC can save lives and can be achieved by screening cirrhotic patients for HCC. Although this strategy is advocated by the major hepatological scientific societies^39,40^, results of this strategy are disappointing due to low adherence. Some experts question the validity of this screening strategy because the incidence of HCC in cirrhosis, despite being the primary risk factor of HCC development, might be lower than generally accepted^5^. A recent prospective French/Belgian cohort of patients with alcoholic-related cirrhosis showed an incidence of HCC of 2.9 per 100 patient-years, and one-year cumulative incidences of 1.8%.

The validity of screening in this situation is an important debate but is not at the heart of this research project. We believe that the question is not whether we should screen these patients or not, but how we can improve the quality, validity, cost-effectiveness and adherence to screening.

Today, the same approach is used in all patients, which leads to disappointingly low adherence rates on the long term^3,41^, for several reasons. Using a more personalised approach might increase the success of HCC screening in cirrhosis. By implementing a precision medicine-based approach, cirrhotic patients should first be stratified according to their expected risk of HCC development. The validation study proposed here provides us with a biomarker that can help answer this question.

The GCT is a serum biomarker that can easily be performed in patients with cirrhosis. The leaf plots (Figure 4) show that this marker has a very high negative predictive value for assessment of risk of HCC development. This information should be used to establish a screening regime tailored to the patients’ needs, preventing unnecessary screening burden in low risk patients. For example, in a patient with a GCT value below 0.2, a yearly screening or even biannual screening regimen could be proposed rather than a 6-monthly screening regimen. This could increase adherence to and cost-effectiveness of screening. In high risk patients, rigorous screening must be adopted. We can add to this the information obtained from serum albumin levels as well.

We are convinced that only the adoption of biomarkers with this ability can move this field forward and usher cirrhosis patient management into the era of personalized medicine.

This study provides an independent validation of the prognostic power of the GCT for this purpose. Also of note is that this biomarker is based on a well-described underlying pathophysiological rationale. An increase in GCT is mainly driven by an increase in bisecting GlcNAc residues, formed by the GnT-III enzyme. This enzyme is increasingly expressed in rat liver dysplastic and malignant nodules during hepatocarcinogenesis^25,41^ and in sera and nodular liver tissue of cirrhotic patients, with and without HCC^26,27^. GCT is specifically increased in cirrhotic patients, but not in patients with earlier stages of liver fibrosis^23^, which supports the hypothesis that GCT increase is related to upregulation of GnT-III in regenerative nodules, the histological hallmark of liver cirrhosis.

Another application of GCT might be to use it as an exclusion criterion for cirrhotic patients in hepatocellular carcinoma-preventing clinical trials. By excluding the patients with the lowest risk of HCC development, the required number of patients for clinical trials could be significantly reduced, saving resources in the organisation of such trials.

Furthermore, the analysis of this biomarker can be transferred to high-throughput automated capillary electrophoresis analysers, already used for serum protein profiling in most modern clinical chemistry laboratories. The GlycoLiverProfile (Helena Biosciences, Newcastle, UK) is the first glycomics-based clinical diagnostic test that has become available to routine clinical chemistry laboratories. This is a last factor that could facilitate the adoption of the biomarker.

## Data Availability

All data produced in the present study are available upon reasonable request to the authors.

## Acknowledgements

This project has received funding from the Grand Challenges Program of VIB. This VIB Program received support from the Flemish Government under the Management Agreement 2017-2021 (VR 2016 2312 Doc.1521/4).

XV received a translational and clinical mandate from Stichting tegen Kanker Belgium.

